# SEVERE COVID-19 IS MARKED BY DYSREGULATED SERUM LEVELS OF CARBOXYPEPTIDASE A3 AND SEROTONIN

**DOI:** 10.1101/2021.02.02.21251020

**Authors:** Rodolfo Soria-Castro, Yatsiri G. Meneses-Preza, Gloria M. Rodríguez López, Sandra Romero-Ramírez, Víctor A. Sosa-Hernandez, Rodrigo Cervantes-Díaz, Alfredo Pérez-Fragoso, José J. Torres-Ruíz, Diana Gómez-Martín, Marcia Campillo-Navarro, Violeta D. Álvarez-Jiménez, Sonia M. Pérez-Tapia, Alma D. Chávez-Blanco, Sergio Estrada-Parra, José L. Maravillas-Montero, Rommel Chacón Salinas

## Abstract

The immune response plays a critical role in the pathophysiology of SARS-CoV-2 infection ranging from protection to tissue damage. This is observed in the development of acute respiratory distress syndrome when elevated levels of inflammatory cytokines are detected. Several cells of the immune response are implied in this dysregulated immune response including innate immune cells and T and B cell lymphocytes. Mast cells are abundant resident cells of the respiratory tract, able to rapidly release different inflammatory mediators following stimulation. Recently, mast cells have been associated with tissue damage during viral infections, but little is known about their role in SARS-CoV-2 infection. In this study we examined the profile of mast cell activation markers in the serum of COVID-19 patients. We noticed that SARS-CoV-2 infected patients showed increased carboxypeptidase A3 (CPA3), and decreased serotonin levels in their serum. CPA3 levels correlated with C-reactive protein, the number of circulating neutrophils and quick SOFA. CPA3 in serum was a good biomarker for identifying severe COVID-19 patients, while serotonin was a good predictor of SARS-CoV-2 infection. In summary, our results show that serum CPA3 and serotonin levels are relevant biomarkers during SARS-CoV-2 infection, suggesting that mast cells are relevant players in the inflammatory response in COVID-19, might represent targets for therapeutic intervention.

## Introduction

Coronavirus disease 2019 (COVID-19) is caused by the SARS-CoV-2 virus, which was identified for the first time in Wuhan, China in late 2019. Since then, the virus has spread across the globe infecting more than 100 million people and causing the death of 2.1 million infected individuals (1).

SARS-CoV-2 infects cells of the mucosa that express the angiotensin-converting enzyme II or ACE2, which is particularly abundant in type II alveolar cells of the lung and also expressed in other tissues (2). The COVID-19 infection has an incubation period of 3-7 days, generating moderate symptoms, such as head and muscle aches, sore throat, nasal congestion, dry cough, fatigue and fever. However, 5-10% of infected patients develop acute respiratory distress syndrome (ARDS), a serious complication that causes respiratory failure leading to high mortality (3).

A relevant factor for the development of ARDS in patients with COVID-19 is the exacerbated immune response triggered by the infection, reflected as a cytokine storm in which different cytokines such as IL-1β, IL-2, IL-4, IL-6, IL-10, IFN-γ and TNF-α are elevated (4). Besides, COVID-19 patients are known to show profound alterations in cell populations associated with the immune response against viruses, such as monocytes, macrophages, neutrophils, NK cells, B lymphocytes, CD8 + T lymphocytes and memory and regulatory CD4 + lymphocytes (5). However, how mast cells (MC) are affected by SARS-CoV-2 is not well understood.

MC are tissue resident leukocytes derived from hematopoietic precursors, distributed throughout the body and abundantly found along the respiratory tract (6). These cells are characterized by presenting many cytoplasmic granules that contain different chemical mediators released after activation. Among the molecules abundant in MC granules are tryptase, carboxypeptidase, chymase, serotonin, histamine and TNF-α. MC also produce other types of inflammatory mediators including prostaglandins, leukotrienes and reactive nitrogen species. In addition, activated mast cells secrete different *de novo* synthesized cytokines and chemokines (7).

MC are well known for their role in mediating allergic reactions, but recent evidence indicates an important role in the innate immune response to different pathogens including viruses, bacteria, protozoa, fungi and nematodes (7). The ability of MC to participate in viral infections is mediated by different Pattern Recognition Receptors, such as TLR-3, −7, RIG-I, MDA-5, etc., which are essential in the innate antiviral response (8). Recently, it was shown that mast cells are associated with tissue damage induced by an excessive inflammatory response during viral infections (9). For instance, in influenza virus infections, the observed lung tissue damage is due to an excessive inflammatory response characterized by the overproduction of cytokines and chemokines or ‘cytokine storm’ (10). In murine models of infection with the H5N1 influenza virus, treatment of mice with ketotifen, an inhibitor of mast cell activation, reduces damage to lung tissue. Further, the combination of ketotifen with oseltamivir, an antiviral drug, significantly increased mice survival (11). On the other hand, it has been observed that mice deficient in MC show less lung damage when infected with influenza A virus, compared to wild-type mice. Interestingly, this effect was associated with a decreased production of TNF-α, CCL2, CCL3, CCL4, CXCL2 and CXCL10, suggesting a crucial role of MC in the ‘cytokine storm’ triggered by influenza infection (12).

Considering that mast cell activation plays a crucial role during the damage induced by viral infections and that cytokine storm is a crucial feature during SARS-CoV-2 infection, this study was conducted to evaluate mast cell activation markers in the serum of patients with COVID-19 and determine if they are associated with disease severity.

## MATERIAL AND METHODS

### Patients and control

Patients with COVID-19 and control group were enrolled at Instituto Nacional de Ciencias Médicas y Nutrición Salvador Zubirán, Mexico City, between March-April 2020. All participants in this study had COVID-19 suggestive symptoms, but only those showing a positive PCR test for SARS-CoV-2 were considered infected, while PCR negative patients were selected as control. Samples from asthmatic, HIV, cancer, autoimmune or pregnant patients were not included in this study. Serum samples were taken upon admission. Demographic and clinical parameters were collected, and laboratory indicators were tested with conventional methods in COVID-19 patients (Supplementary Table 1). Disease severity was classified as Mild/Moderate when patients showed fever, signs of airway disease, with or without a tomographic image indicating pneumonia. Severe COVID-19 patients showed either respiratory failure, an increased respiratory rate (>30 bpm), decreased oxygen saturation at rest (<92%) or decreased PaO_2_/FiO_2_ (<300 mmHg). This study was approved by the Research and Ethics Committee of the Instituto Nacional de Ciencias Médicas y Nutrición Salvador Zubirán (Ref. 3341). All protocols were in accordance with the Declaration of Helsinki. All participants of this study signed an informed consent form to participate in this study.

### Assessment of mast cell activation markers

Patient serum was analyzed by ELISA with commercial kits for histamine (Cat. RE59221, IBL International, Germany), human carboxypeptidase A3 (Cat. OKCD01671, Aviva Systems Biology, USA), serotonin (Cat. ab133053, Abcam, UK) and heparin (Cat. abx 258893, Abbexa, USA), according to the manufacturer’s instructions. Nitric oxide was evaluated by a colorimetric method that measures the levels of the breakdown product NO_2_^-^ (Griess Reagent System, Cat. G2930, Promega, USA).

### IL-6 quantification

IL-6 was evaluated with the Milliplex Map Human Cytokine/Chemokine Bead Panel-Premixed 29 Plex (Cat. HCYTMAG-60K-PX29, Millipore, USA) following manufacturer’s instructions.

### Statistical analysis

All statistical analyses were performed with SigmaPlot software version 14.0 (Systat Software, San Jose, CA, USA). Data normality was assessed by Kolmogorov-Smirnov with Lilliefors correction. Data are shown as mean or median ± range, as appropriate. For comparisons between two groups, Student’s t-test or Mann–Whitney rank sum test with Yates correction were used. For comparisons of three groups, one way-analysis or variance (ANOVA) with Student-Newman-Keuls (SNK) post-hoc or Kruskal-Wallis test followed by a Dunn’s post-hoc test were used. Results were considered significant at a p value <0.05. For correlations of two variables, Spearman Rank Order Correlation was used. The r values for each of the correlations were plotted in a bubble chart generated with Microsoft Excel 2019 (https://office.microsoft.com/excel). Receiver operating characteristic (ROC) curves were generated to find the accuracy of biomarkers to distinguish infected individuals and the severity of COVID19.

## RESULTS AND DISCUSSION

Several MC-derived biomarkers are used to diagnose and predict outcomes in allergic and infectious diseases. Among them, histamine, heparin, carboxypeptidase A3 (CPA3), serotonin, and nitric oxide have been studied (13, 14). To address whether these markers are affected during COVID-19, serum samples were collected from patients admitted at a tertiary care center in Mexico city. Demographic and clinical characteristics of patients are shown in supplementary table 1. We noticed that COVID-19 patients have increased levels of CPA3 in serum when compared to the control group (Figure 1A). Interestingly, serotonin showed decreased levels in serum of SARS-CoV-2 infected patients compared to those not-infected (Figure 1B). Serum levels of histamine, heparin, and nitric oxide were not affected in COVID-19 patients (Supplementary Figure 1). Previous reports indicate that different immune markers are differentially expressed during severe COVID-19 (15). Therefore, we investigated whether mast cell-derived biomarkers were altered depending on COVID-19 severity. We observed that patients with severe disease showed increased levels of CPA3 when compared to those with mild/moderate disease, or individuals in the control group (Figure 1C). Serotonin levels were decreased in severe COVID-19 (Figure 1D). No other mast cell-associated biomarker evaluated showed significant difference in patients with severe COVID-19, compared to patients with mild/moderate symptoms or controls (Supplementary Figure 2).

**Figure 1.**
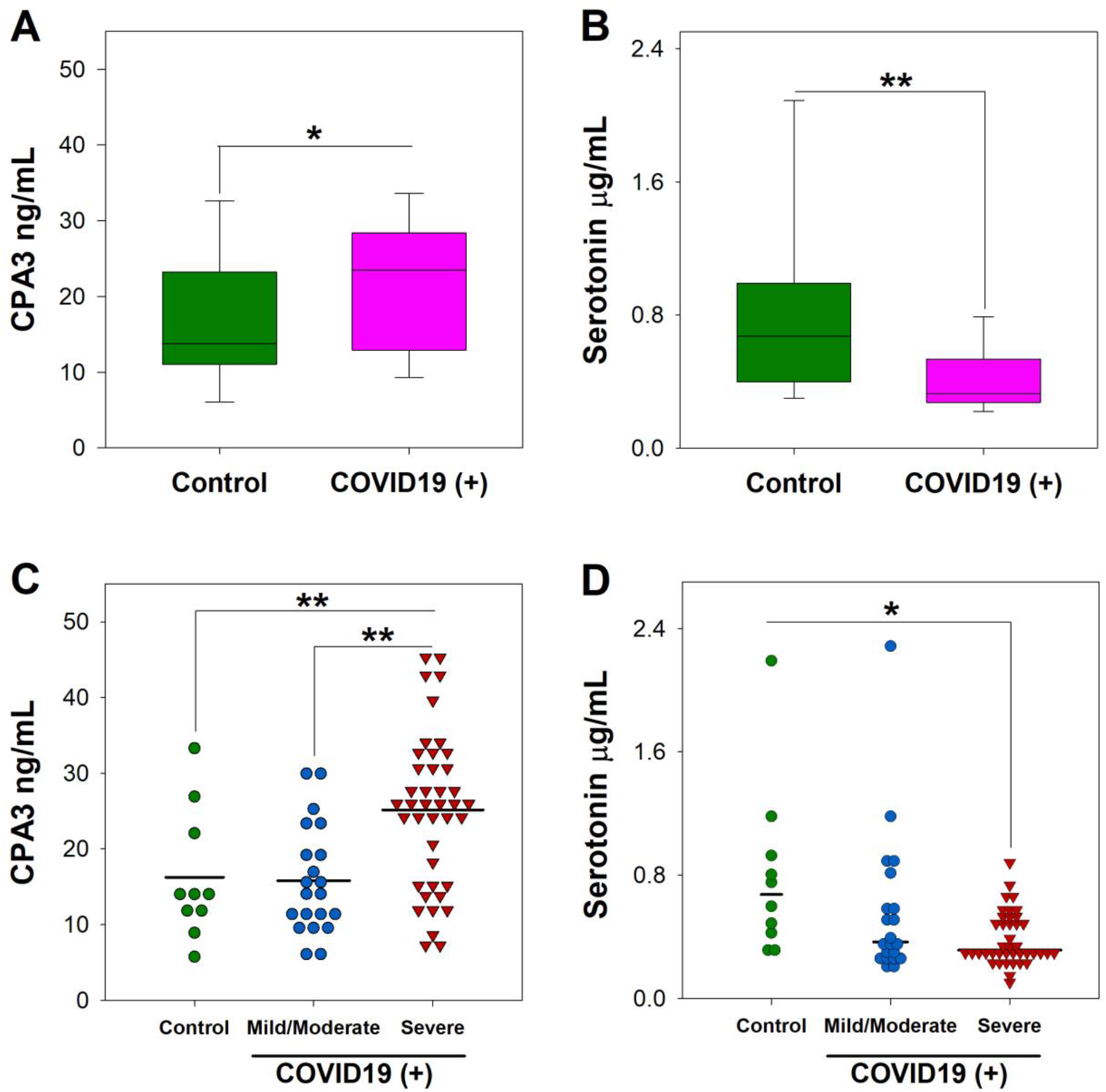
Serotonin and carboxypeptidase A3 are altered in COVID-19 patients. The serum concentration of A, C) Carboxypeptidase A3 (CPA3), and B, D) Serotonin were measured upon patient admission by ELISA. Data from 21 patients with mild/moderate disease, 41 patients with severe COVID-19, and 10 control individuals is shown. Data are presented as mean or median ± range, as appropriate. *p<0.05; **p< 0.01. Student’s t test (A) Mann-Whitney test (B), one-way ANOVA test (C) and Kruskal-Wallis test (D)

Because CPA3 and serotonin levels were altered in severe COVID-19 patients, we next assessed their correlation with clinical and laboratory parameters. A matrix with Spearman’s r coefficient values and representing correlation showed that CPA3 had a stronger correlation with an increased number of clinical and laboratory parameters in comparison with serotonin (Figure 2A). Furthermore, CPA3 showed significative positive correlation with inflammation associated markers as circulating neutrophils (r=0.291, p=0.0447) (Figure 2B) and C-reactive protein (r=0.390, p=0.00703) (Figure 2D). Remarkably, CPA3 also associated with disease severity score quick Sepsis-related Organ Failure Assesment (qSOFA), (r=0.335, p=0.00862) (Figure 2C).

**Figure 2.**
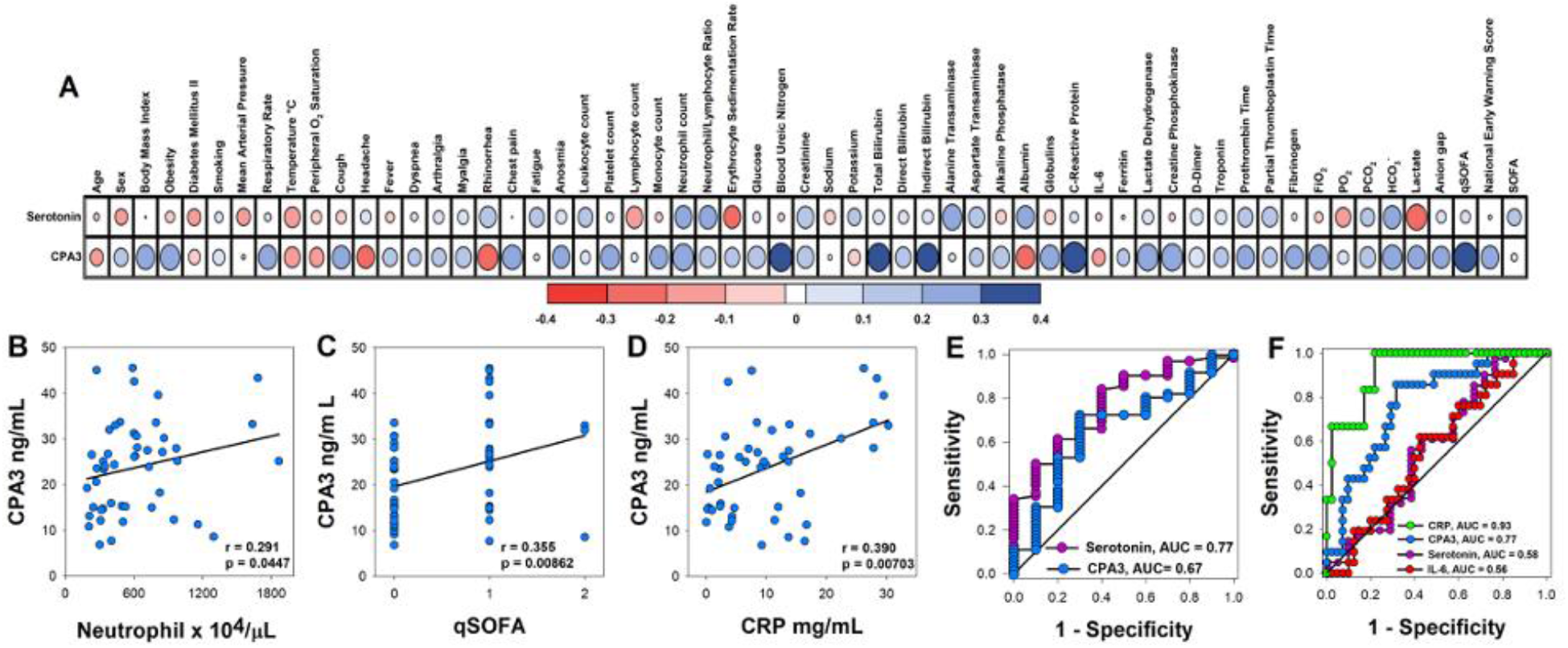
Serum levels of carboxypeptidase A3 correlates with clinical parameters of disease severity in COVID-19 patients. A) Correlation matrix representing correlation carboxypeptidase A3 or serotonin serum levels with clinical and laboratory parameters used to determine COVID-19 severity. Spearman’s coefficient value r is used as correlation descriptor, and the size of each circle symbolizes correlation strength (color scale of red and blue indicates negative or positive correlation, respectively). Correlation between serum concentration of carboxypeptidase A3 and B) blood neutrophils, C) qSOFA, D) C-reactive protein. Value of Sperman’s correlation (r) and significant p values (p<0.05) are shown. E) Receiver-operator characteristics (ROC) curves of carboxypeptidase A3 (CPA3) and serotonin serum levels for the prediction of SARS-CoV-2 infection. F) ROC curve of C-reactive protein (CRP), carboxypeptidase A3 (CPA3), serotonin and interleukin-6 (IL-6) in the prediction of COVID-19 severity.

Finally, to evaluate the potential clinical utility of serotonin and CPA3, receiver-operator characteristics (ROC) curves were performed to distinguish SARS-CoV-2 infection. Serotonin showed an acceptable AUC (Area Under the Curve) values that allow to distinguish between infected from non-infected individuals (AUC 0.77) (Figure 2E). When ROC curves were analyzed to differentiate between mild/moderate and severe COVID-19 patients, CPA3 (AUC 0.77) was more reliable than serotonin (AUC 0.58) or IL-6 (AUC 0.56) to detect patients with severe disease. This was close to the predictive values seen for C-reactive protein (AUC 0.93) (Figure 2F). As a whole, these results suggest a relationship between mast cell activation, as reflected by CPA3 levels in serum, and severe COVID-19.

To the best of our knowledge, this is the first evidence of alteration in serum serotonin and CPA3 levels during COVID-19. CPA3 is abundantly expressed in human lungs (16), with MC being the main cell source (Supplementary Figure 3). CPA3 is an enzyme that cleaves C-terminal amino acid residues from proteins and is an abundant protein in MC granules (16 μg CPA3 per 10^6^ MC). This enzyme is released after cell degranulation and is associated with allergic pathologies of the respiratory tract (17). Identified substrates for CPA3 include neurotensin, kinetensin, neuromedin N, angiotensin I and endothelin-1, which is associated with pulmonary fibrosis (18), a sequela observed in COVID-19 patients (19). Interestingly, we noticed that CPA3 correlated with clinical parameters associated with systemic inflammation during COVID-19. Remarkably, CPA3 correlated with circulating neutrophils and CPR, which are associated with an exacerbated inflammatory response during COVID-19 (15). Previous studies have noticed the importance of MC activation for the recruitment of neutrophils to sites of infection (20). Furthermore, this increase in tissue neutrophil is proposed as one mechanism of tissue damage and organ failure during COVID-19 (21). Our results are in agreement with a recent histologic study, where an increased number of MC in the lungs of COVID-19 patients was observed (22). These results suggest an important role of MC in SARS-CoV-2 infection, but further work is needed to understand the mechanisms involved.

The second marker that was modified during SARS-CoV-2 infection was serotonin. Traditionally serotonin is considered as an important neurotransmitter regulating several neuronal activities in the central nervous system. However, recent evidence indicates that systemic serotonin distributed by the blood plays a more complex function in the organism. Blood serotonin is produced by different cell lineages, including MC, but is mainly produced by enterochromaffin cells of the intestine. Once in the blood, serotonin levels are primarily regulated by platelets which capture and store in dense granules which are secreted after cell activation (23). Previous studies have shown that infections can decrease, increase or maintain serotonin levels in blood (24-26). For instance, in dengue infection decreased levels of serum serotonin were associated with disease severity. Moreover, a correlation is observed between decreased serotonin and thrombocytopenia, a clinical feature of severe dengue, associating decreased serum serotonin with decreased numbers of platelets (24, 26). However, we did not observe a difference in platelet count in COVID-19 patients (Supplementary Table 1), implying other mechanisms could be involved in this decrease. A recent report noticed that platelet numbers are not affected in COVID-19, but their gene expression profile is different when compared with those from healthy individuals (27). Because platelets usually introduce serotonin from blood through a transporter (SERT or SLC6A4) (28), an overexpression of this protein could explain the decrease of serotonin in blood.

However, Manne *et a*l showed that SERT is not modified in platelets from COVID-19 patients (27). Another explanation could be related to an alteration in enterochromaffin cells functions by SARS-CoV-2, a phenomenon that is observed in other viral infections (29), or to increased serotonin degradation by monoamine oxidase (23), which is overexpressed by platelets in COVID-19 patients (27). How this serotonin decrease can affect the pathophysiology of SARS-CoV-2 infection is unknow; however among the diverse functions of blood serotonin, one involves the modulation of immune response. Several cells of the immune system express serotonin receptors, including those that participate in innate and adaptive immune response (23). Activation of serotonin receptor 5-H2TA diminishes inflammation induced by TNF-α (30). Furthermore, serotonin downmodulates IL-6 and TNF-α production by macrophages and lymphocytes (31), suggesting the importance of serotonin in regulating exacerbated inflammation. In conclusion, our results demonstrate that serum levels of CPA3 and serotonin are affected during SARS-CoV-2 infection and can be considered as biomarkers during COVID-19. We suggest that mast cells play an important role in this disease and that these cells could be considered targets for therapeutic intervention.

## Data Availability

The data supporting the conclusions of this article will be made available by the authors upon reasonable request.

## AUTORSHIP

RS-C, YGM-P and GMR-L contributed equally to this work, designed, performed experiments, analyzed data and drafted the manuscript. SR-R, VAS-H, RC-D, AP-F, JJT-R, DG-M assisted in processing and preservation of patient samples, collected patient data, generated, and organized clinical database. MC-N, VDA-J, SMP-T, ADC-B, SE-P, JLM-M and RC-S analyzed, interpreted data, and drafted the manuscript. JLM-M and RC-S designed, supervised the study and obtained funding. All authors critically revised and approved the final version of this manuscript.

## ACKNOWLEDGMENTS

We want to thank Dr Eduardo Ramírez San Juan for his invaluable advice for the statistical analysis of our study, Fabián Flores-Borja for critical reading of the manuscript, and Araceli Olvera G. for secretarial assistance. This study was supported by CONACyT (F0005-2020-01-313252) to JLM-M, and (F0005-2020-01-312326) to RC-S, and Secretaría de Investigación y Posgrado del IPN (SIP-IPN).

## DISCLOSURE

The authors declare no conflicts of interest.

## Notes

### Competing Interest Statement

The authors have declared no competing interest.

### Funding Statement

This study was supported by CONACyT (F0005-2020-01-313252) to JLM-M, and (F0005-2020-01-312326) to RC-S, and Secretaria de Investigacion y Posgrado del IPN (SIP-IPN).

### Author Declarations

This study was approved by the Research and Ethics Committee of the Instituto Nacional de Ciencias Medicas y Nutricion Salvador Zubiran (Ref. 3341). All protocols were in accordance with the Declaration of Helsinki. All participants of this study signed an informed consent form to participate in this study.

