## Supplemental Table 1, Figures 1 and 2 for "SEVERE COVID-19 IS MARKED BY DYSREGULATED SERUM LEVELS OF CARBOXYPEPTIDASE A3 AND SEROTONIN"

**Article type:** Brief conclusive report

**Key words:** Mast cell, carboxypeptidase A3, serotonin, SARS-CoV-2, COVID-19.

1. Departamento de Inmunología, Escuela Nacional de Ciencias Biológicas, Instituto Politécnico Nacional, ENCB-IPN. Mexico City, Mexico.

2. Red de Apoyo a la Investigación, Universidad Nacional Autónoma de México e Instituto Nacional de Ciencias Médicas y Nutrición Salvador Zubirán, Mexico city, Mexico.

3. Facultad de Medicina, Universidad Nacional Autónoma de México, Mexico city, Mexico.

4. Departamento de Biomedicina Molecular, Centro de Investigación y de Estudios Avanzados del Instituto Politécnico Nacional, Mexico City, Mexico.

5. Departamento de Inmunología y Reumatología, Instituto Nacional de Ciencias Médicas y Nutrición Salvador Zubirán, Mexico City, Mexico.

6. Departamento de Atención Institucional Continua y Urgencias, Instituto Nacional de Ciencias Médicas y Nutrición Salvador Zubirán, Mexico City, Mexico.

7. Research Coordination, Centro Médico Nacional 20 de Noviembre, ISSSTE, Mexico City, Mexico

8. Lab. de Biología Molecular y Bioseguridad Nivel 3. Centro Médico Naval-SEMAR, Mexico city, Mexico

9. Unidad de Desarrollo e Investigación en Bioprocesos (UDIBI), Escuela Nacional de Ciencias Biológicas, Instituto Politécnico Nacional, ENCB-IPN. Mexico City, Mexico

10. División de Ciencia Básica, Instituto Nacional de Cancerología (INCan). Mexico City, Mexico

‡ These authors have contributed equally to this work

* Corresponding authors:

José L. Maravillas-Montero, Ph.D.

Rommel Chacón-Salinas, Ph.D.

### Abstract

The immune response plays a critical role in the pathophysiology of SARS-CoV-2 infection ranging from protection to tissue damage. This is observed in the development of acute respiratory distress syndrome when elevated levels of inflammatory cytokines are detected. Several cells of the immune response are implied in this dysregulated immune response including innate immune cells and T and B cell lymphocytes. Mast cells are abundant resident cells of the respiratory tract, able to rapidly release different inflammatory mediators following stimulation. Recently, mast cells have been associated with tissue damage during viral infections, but little is known about their role in SARS-CoV-2 infection. In this study we examined the profile of mast cell activation markers in the serum of COVID-19 patients. We noticed that SARS-CoV-2 infected patients showed increased carboxypeptidase A3 (CPA3), and decreased serotonin levels in their serum. CPA3 levels correlated with C-reactive protein, the number of circulating neutrophils and quick SOFA. CPA3 in serum was a good biomarker for identifying severe COVID-19 patients, while serotonin was a good predictor of SARS-CoV-2 infection. In summary, our results show that serum CPA3 and serotonin levels are relevant biomarkers during SARS-CoV-2 infection, suggesting that mast cells are relevant players in the inflammatory response in COVID-19, might represent targets for therapeutic intervention.

**Supplementary Table 1. Demographics and clinical characteristics of COVID-19 patients and control group.**

| Demographics | Control | Mild/Moderate | Severe |
|---|---|---|---|
| n | 10 | 21 | 41 |
| Age § | 34 ± 3.6 | 39 ± 2.5 | 48 ± 2.2 |
| Famale (%) | 4/10 (40%) | 11/21 (52.4%) | 16/42 (38%) |
| Male (%) | 6/10 (60%) | 10/21 (47.6%) | 26/42 (62%) |
| **Co-morbidities** | | | |
| Obesity (%) | 1/10 (10%) | 3/21 (14.3%) | 18/41 (43.9%) |
| Diabetes Mellitus (%) | 0/10 (0%) | 1/21 (4.8%) | 8/41 (19.5%) |
| Hypertension (%) | 0/10 (0%) | 0/21 (0%) | 16/41 (39%) |
| Cardiopathy (%) | 0/10 (0%) | 0/21 (0%) | 4/40 (10%) |
| Cerebrovascular disease (%) | 0/10 (0%) | 0/21 (0%) | 1/41 (2.4%) |
| Chronic Kidney Disease (%) | 0/10 (0%) | 1/21 (4.8%) | 0/41 (0%) |
| Liver disease (%) | 1/10 (10%) | 0/21 (0%) | 2/41 (4.9%) |
| Smoker (%) | 2/10 (20%) | 1/21 (4.8%) | 5/41 (12.2%) |
| **Symptoms** | | | |
| Days of symptoms † | | 3 (2-7.25) | 8 (6-11.25) |
| Fever (%) | 9/10 (90%) | 16/21 (76.2%) | 37/41 (90.2%) |
| Cough (%) | 7/10 (70%) | 17/21 (81%) | 34/41 (82.9%) |
| Headache (%) | 8/10 (80%) | 17/21 (81%) | 22/41 (53.7%) |
| Dyspnea (%) | 6/10 (60%) | 5/21 (23.8%) | 31/41 (75.6%) |
| Arthralgia (%) | 4/10 (40%) | 12/21 (57.1%) | 27/41 (65.9%) |
| Myalgia (%) | 4/10 (40%) | 13/21 (61.9%) | 29/41 (70.7%) |
| Odynophagia (%) | 3/10 (30%) | 4/21 (19%) | 14/41 (34.1%) |
| Rhinorrhea (%) | 3/10 (30%) | 9/21 (42.9%) | 9/41 (22%) |
| Conjunctivitis (%) | 2/10 (20%) | 3/21 (14.3%) | 2/41 (4.9%) |
| Chest pain (%) | 4/10 (40%) | 1/21 (4.8%) | 7/41 (17.1%) |
| Vomit (%) | 3/10 (30%) | 1/21 (4.8%) | 3/41 (7.3%) |
| Diarrhea (%) | 3/10 (30%) | 3/21 (14.3%) | 8/41 (19.5%) |
| **Vital signs** | | | |
| Respiratory rate (breaths/minute) † | 20 (17.75-23) | 18 (17-24) | 24 (20-29.5) |
| Oxygen saturation (%)† | 95 (92.25-97.25) | 95 (93.5-95.5) | 88 (85-91.5) |
| Hear rate (beats/minute) § | 104 ± 6.7 | 99 ± 4.4 | 105 ± 2.6 |
| Mean arterial pressure (mmHg) § | 85 ± 3.3 | 93 ± 2.5 | 93 ± 2.1 |
| **Measures of illness severity** | | | |
| SOFA= 0 | | 4/4 (100%) | 5/27 (18.5%) |
| SOFA= 1 | | | 19/27 (70.4%) |

| | | |
|---|---|---|
| SOFA >1 (median= 2) | | 3/27 (11.1%) |
| qSOFA= 0 | 15/15 (100%) | 12/38 (31.6%) |
| qSOFA= 1 | | 23/38 (60.5%) |
| qSOFA= 2 | | 3/38 (7.9%) |
| NEWS [Median (Q1-Q3)] † | 3 (0.3-4.8) | 6 (4-9) |
| PSI/PORT § | | 61 ± 4.3 |
| **Laboratory values** | | |
| White cell count ($x10^9$/L)† | 6.7 (5.2-10.7) | 7.2 (5.1-10.2) |
| Lymphocytes ($x10^9$/L) § | 0.97 ± 0.2 | 0.86 ± 0.06 |
| Monocytes ($x10^9$/L) § | 0.35 ± 0.09 | 0.5 ± 0.04 |
| Neutrophils ($x10^9$/L) † | 3.3 (2.1-4.0) | 6.0 (3.6-8.4) |
| Neutrophils/Lymphocytes † | 2.3 (1.7-2.9) | 7.7 (4.2-12.5) |
| Platelets ($x10^9$/L) § | 218 ± 21.8 | 221.1 ± 11.8 |
| Hemoglobin (g/dL) § | 16.4 ± 0.6 | 15.4 ± 0.2 |
| ESR (mm/hour) † | | 17 (10-41) |
| Blood glucose (mg/dL) † | 91 (85-100) | 111 (102.5-124.5) |
| BUN (mg/dL) † | 10.5 (10.2-12.5) | 12.8 (10-19.7) |
| Creatinine (mg/dL) † | 0.9 (0.7-1.0) | 0.9 (0.8-1.1) |
| Total bilirubin (mg/dL) § | 0.6 ± 0.1 | 0.6 ± 0.03 |
| Direct bilirubin (mg/dL) † | 0.1 (0.09-0.2) | 0.2 (0.1-0.2) |
| Indirect bilirubin (mg/dL) † | 0.4 (0.2-0.5) | 0.5 (0.3-0.6) |
| Alanine aminotransferase level (IU/L) † | 26 (11.7-57) | 38.6 (22.5-59.3) |
| Aspartate aminotransferase level (IU/L) † | 26 (17.7-36) | 47 (26.7-63.7) |
| Alkaline phosphatase (IU/L) § | 88 ± 5.7 | 95.5 ± 6.6 |
| Lactate dehydrogenase (IU/L) † | 176 (136.8-229.5) | 315 (268.5-439.8) |
| Creatine kinase (IU/L) † | 61.5 (49-113.8) | 105.5 (50.5-239.5) |
| **Gasometry values** | | |
| $FiO_2$ (%)† | 21 (21-21) | 21 (21-30) |
| Arterial pH † | 7.4 (7.4-7.6) | 7.46 (7.4-7.5) |
| $PaO_2$ (mmHg) † | 66.6 (61.1-69.6) | 63.2 (52-78.1) |
| $PCO_2$ (mmHg) † | 31.7 (20.1-34.7) | 30.7 (28.2-32.5) |
| Lactate (mmol/L) † | 0.9 (0.5-1.0) | 1.2 (1.0-1.6) |
| $HCO_3^-$ (mmol/L) † | 19.6 (18.8-23.2) | 21.8 (19.9-22.7) |
| $PaO_2/FiO_2$† | 317 (291-331) | 248 (223-306) |

† [Median (Q1-Q3)], §(Mean ± SEM), SOFA, Sequential Organ Failure Assessment; qSOFA, quick SOFA; NEWS, National Early Warning Score; PSI/PORT, Pneumonia Severity Index; ESR, Erythrocyte Sedimentation Rate; BUN, Blood Urea Nitrogen; PT, Prothrombin Time; PTT, Partial Thromboplastin Time; $FiO_2$, Fraction of Inspired Oxygen; $PaO_2$, Partial Pressure of Oxygen; $PCO_2$, Partial Pressure of Carbon Dioxide

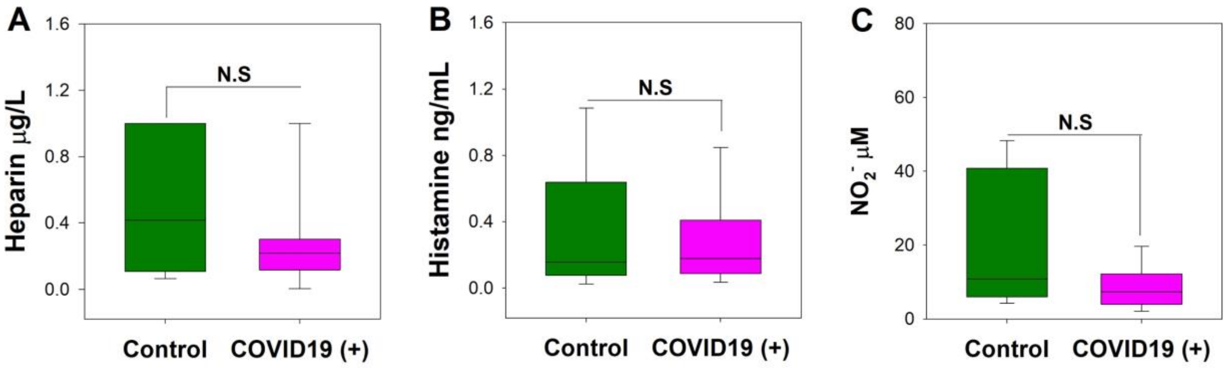


**Supplementary Figure 1. Serum levels of heparin, histamine, and nitrites are not altered in severe COVID-19 patients**. Concentration of A) Heparin, B) Histamine and C) Nitrites was evaluated in serum of COVID-19 patients. Data from 21 patients with mild/moderate COVID-19, 41 severe disease and 10 control individuals are shown as median ± range. Mann-Whitney test.

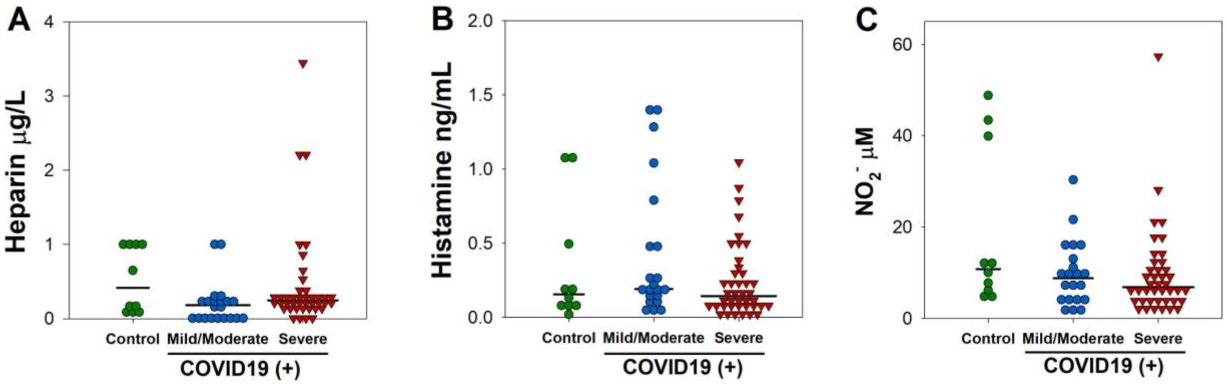


**Supplementary Figure 2. Serum levels of heparin, histamine, and nitrites are not altered in severe COVID-19 patients**. Concentrations of A) Heparin, B) Histamine, and C) Nitrites was evaluated in serum of COVID-19 patients. Data from 21 patients with mild/moderate COVID-19, 41 severe disease and 10 control individuals are shown as median ± range. Kruskal-Wallis test.

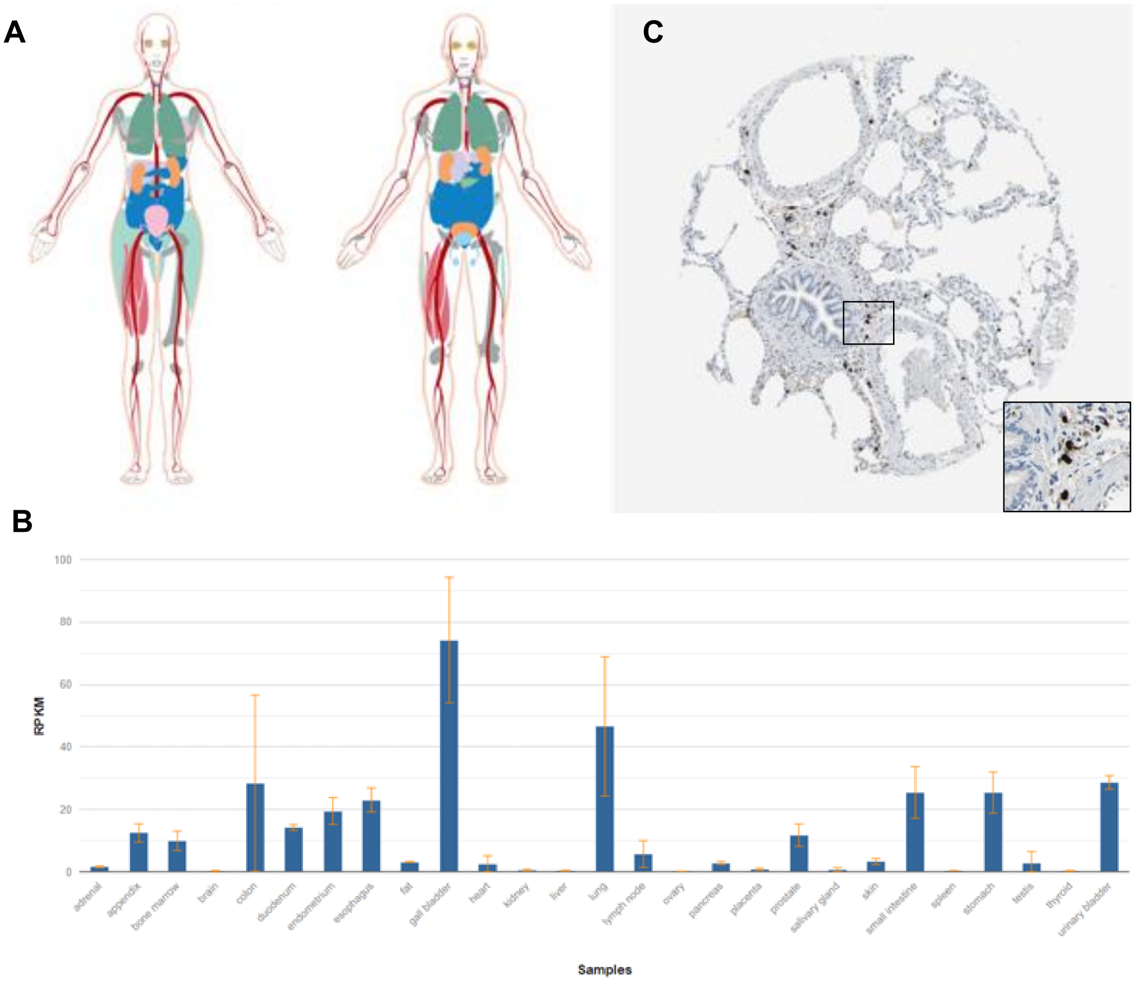


**Supplementary Figure 3. Protein and mRNA CPA3 expression in different human organs. A)** Human CPA3 protein expression in different organs and tissues **B)** mRNA CPA3 expression in different human organs and tissues. **C)** Immunohistochemistry image of CPA3 expression in human lung. The insert image represents a high magnification of the selected zone. RPKM (Reads Per Kilobase Million). Images A,C) were obtained from Human Protein Atlas (16), located at: https://www.proteinatlas.org/ENSG00000163751-CPA3/tissue) and image B) from HPA RNA-seq normal tissues, located at: https://www.ncbi.nlm.nih.gov/gene/1359#gene-expression
